# Integration of stepped care for perinatal mood and anxiety disorders among women attending maternal and child health clinics in Kenya: Protocol for a cluster randomized controlled trial

**DOI:** 10.64898/2026.05.06.26352574

**Authors:** Agnes Karingo Karume, Nancy Ngumbau, Linnet Ongeri, Asterico Neema, Anne Kaggiah, David Owaga, Catherine Maina, Clinton Ndambuki, Yuwei Wang, Lincoln Pothan, Alexandra Rose, Anna Larsen, Sarah Masyuko, Barbra A Richardson, Anjuli D Wagner, Amritha Bhat, Keshet Ronen, John Kinuthia

## Abstract

**Background:** Perinatal mood and anxiety disorders (PMAD) cause substantial morbidity and mortality globally. Kenya, like many low- and middle-income countries, experiences severe shortage of specialized mental healthcare workers and poor coverage of screening and management for PMAD. Provision of screening and treatment by lay and non-specialist providers in the context of routine maternal child health services may enhance access to mental health services and improve outcomes.

**Methods and analysis:** In a hybrid type II cluster randomized clinical trial in 20 facilities in Western Kenya, we will evaluate the clinical, service delivery, and implementation outcomes of a stepped-care model integrating: 1) screening of all pregnant women for depression and anxiety symptoms, 2) Problem Management Plus (PM+) counseling delivered by trained lay providers, and 3) telepsychiatry for women with severe symptoms, suicidality, or inadequate response to PM+. The intervention package also includes a bundle of implementation strategies such as audit and feedback, process mapping, health educational health talks, and procurement of antidepressants for all sites. In addition, intervention sites will receive provider training, structured supervision and financial compensation for providers.

Facilities will be randomized 1:1 to intervention and enhanced standard of care (basic screening and referral to specialist care), using restricted randomization. We will enroll a total of 2,970 women. Women will be eligible if they are pregnant and ≥20 weeks’ gestation, attending antenatal care at the facility, ≥14 years old, and screen positive for PMAD symptoms (Patient Health Questionnaire [PHQ]-2≥3 and/or Generalized Anxiety Disorder-[GAD]-2≥3). Assessments will be conducted at enrollment (pregnancy), 6 weeks, 14 weeks, and 6 months postpartum among participants in both study arms to align with routine well-baby visits.

Primary outcomes are PMAD symptoms (PHQ-9 and GAD-7 score) change at 14 weeks postpartum using an intent-to-treat analysis. Secondary outcomes include quality of life at 14 weeks postpartum, PM+ mechanism of action, and adverse pregnancy outcomes at 6 weeks postpartum. Using mixed methods, we will evaluate acceptability and fidelity of the intervention and compare service delivery and other implementation outcomes (penetration, efficiency, equity, cost) across arms and alongside multilevel drivers of implementation success.

**Ethics and dissemination:** The study protocol was approved by Kenyatta National Hospital/University of Nairobi (P425/04/2023) and the University of Washington (STUDY00017933). All participants will provide written informed consent. Findings will be published in peer-reviewed journals and international conferences.

**Trial registration number:** NCT06456307

**Strengths and limitations of this study:** Strengths include:

- This is among the first studies to evaluate a stepped care intervention for perinatal mental health that has been integrated into routine public maternal child health services in a low-resource setting
- This is among the first studies to evaluate telepsychiatry as part of an integrated care model in a low-resource setting.
- This study combines 3 evidence-based and resource-appropriate interventions, that have the potential to be scalable in this context
- The study will generate not only clinical effectiveness data but also data on implementation outcomes in the context of service provision by routine providers, which will enable holistic evaluation of the intervention’s potential
- This is a large randomized controlled trial conducted in a mix of rural and urban clinics in Kenya, increasing generalizability across geographical contexts

Limitations include:

- PM+ is provided in-person and transportation barriers may prevent clients’ consistent attendance
- The implementation strategies used include financial support for providers and essential medications, limiting generalizability

## Introduction

Perinatal mood and anxiety disorders (PMAD), defined as depression and anxiety during pregnancy or up to one year postpartum, is a global public health challenge that negatively impacts the health of women and their children. It is associated with adverse pregnancy outcomes (preterm delivery, low birth weight, pregnancy loss); negative childhood outcomes (insecure infant attachment, poor neurocognitive development, poor growth and physical development, and emotional-behavioral difficulties) and maternal morbidity and mortality (by suicide)^1–4^. Perinatal women in low- and middle-income countries (LMICs) experience a disproportionate burden of PMAD with a prevalence of 20% and higher^5–7^ due to higher occurrence of risk factors such as gender inequality, teenage pregnancies, poverty and intimate partner violence^8,9^. Despite the high burden of PMAD in LMICs, strategies to address this public health issue within resource-limited settings remain under-emphasized and under-researched.

Like many LMICs, Kenya faces a severe shortage of specialized mental healthcare workers (HCWs), with only one psychiatrist per 500,000 residents compared to the recommended ratio of one per 30,000^10,11^. This shortage has resulted in limited coverage of screening and management for PMAD. Consequently, more than 75% of individuals who need mental health services do not receive them, while those who do, must travel long distances and endure lengthy waits to access specialist care^8,12,13^.The World Health Organization (WHO) and Kenya’s national mental health strategy have prioritized the integration of mental health care in primary care settings^14,15^. Maternal child health (MCH) care and prevention of mother-to-child transmission of HIV (PMTCT) clinics are widely attended in sub-Saharan Africa. In Kenya, over 95% of all pregnant women receive at least one antenatal care (ANC) service from a skilled provider^16^. These clinics therefore offer a highly accessible point for integration of perinatal mental healthcare services, in line with the Kenya Mental Health Action Plan 2021-2025’s goal of expanding access to mental health services via MCH and HIV care programs^15^.

To ensure optimal and sustainable integration of perinatal mental healthcare within the existing MCH and PMTCT clinics, resource-appropriate service delivery models should be used and tailored implementation strategies should be developed and tested. Several evidence-based interventions (EBIs) for identification and management of PMAD are available that can be delivered in resource-limited settings by lay workers and non-specialist HCWs^17–20^. The Mental Health Gap Action (mhGAP) Programme developed by WHO provides evidence-based guidance on approaches to integrate mental health screening and care using the resources available within an existing healthcare system^21,22^. In this trial, we integrate three evidence-based interventions into a stepped care model. The first step is screening of all pregnant women using Patient Health Questionnaire-2 (PHQ-2) and Generalized Anxiety Disorder-2 (GAD-2) by lay providers (HIV Testing Service [HTS] providers or mentor mothers), followed by screening by nurses using PHQ-9/GAD-7 for women identified as symptomatic ^22^. The second step is lay-delivered Problem Management Plus (PM+), an evidence-based low-intensity psychological intervention endorsed by WHO, designed to address common mental health problems, such as depression, anxiety, and stress, particularly in low-resource settings^23,24^. The third step is higher-intensity treatment through telepsychiatry for women who do not respond to PM+, experience suicidality, and/or have severe symptoms^25–27^.

We propose to evaluate the clinical and implementation outcomes of this stepped care model for perinatal mental health integrated into MCH and PMTCT clinics. We will also test a bundle of implementation strategies, developed through formative work with stakeholders: essential medicine provision (sertraline) to ensure medication availability, audit and feedback, process mapping and educational health talks for all the sites, as well as ongoing provider training, structured supervision and provider financial compensation for intervention sites only. We hypothesize that integration of these interventions together with implementation strategies into routine perinatal services will reduce PMAD symptoms and adverse perinatal outcomes, as well as improve acceptability, penetration, and equity of care for vital mental health services among perinatal women in Kenya.

## Methods

### Study design

The Integrated Perinatal Mental Health (IPMH) study is non-blinded cluster-randomized controlled superiority trial (cRCT) comparing two parallel arms. Intervention facilities provide a stepped care model of mental health screening for all pregnant women, PM+ counseling, and telepsychiatry. Control facilities provide enhanced standard of care: initial PMAD screening (PHQ-2/GAD-2 screening by HTS providers and mentor mothers) followed by standard of care referral (same-day handoff to an on-site counselor/psychologist when available; otherwise, a written referral to the county psychiatry clinic). This is a hybrid type II effectiveness-implementation trial^28^ (Figure 1).

**Figure 1.**
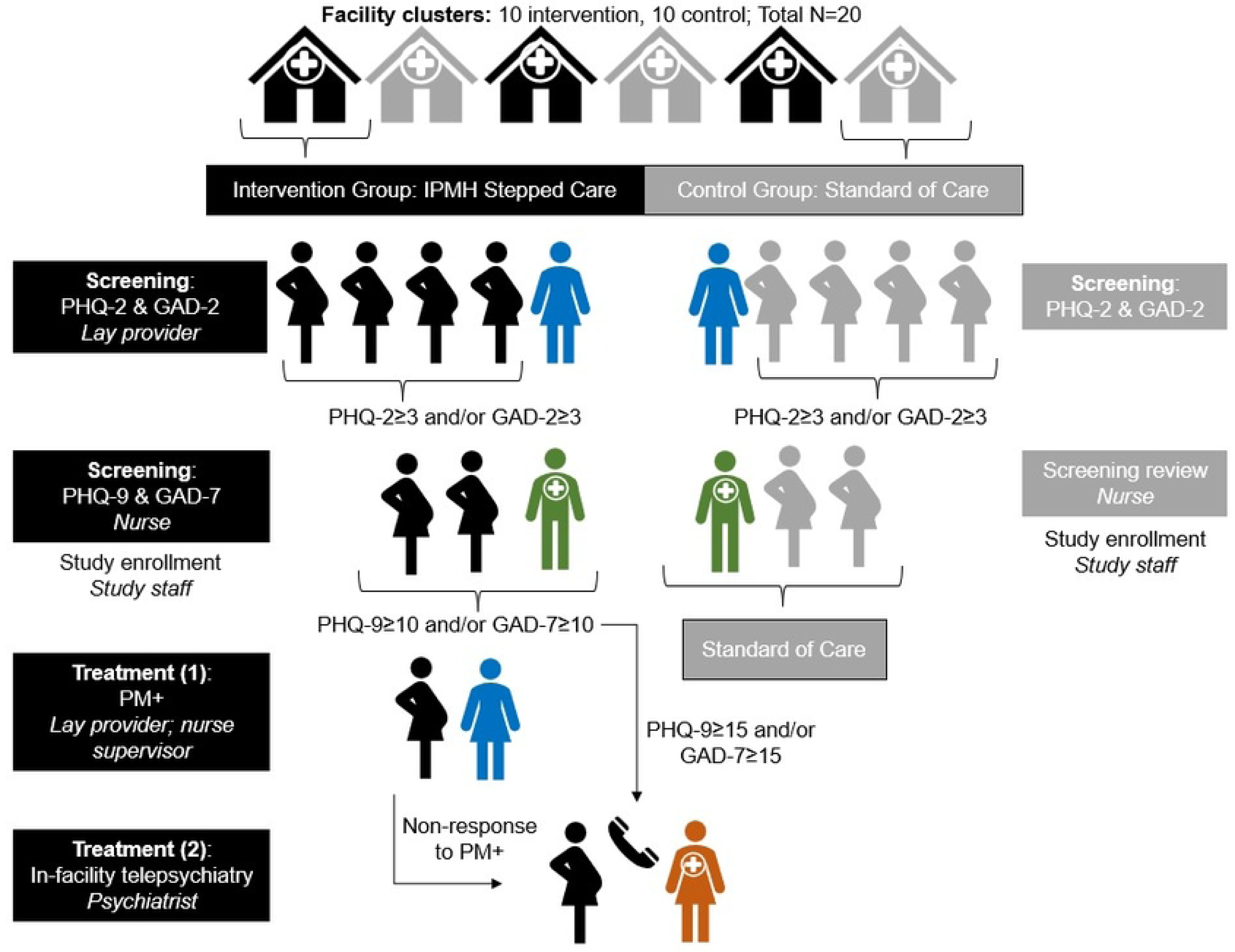
Schedule of interventions and assessments

### Study setting and site selection

The study is conducted in 20 facilities in Western Kenya. Study sites were selected in Kisumu, Siaya and Homa Bay Counties, based on ANC volume (≥20 new ANC clients per month), and including a diversity of facility characteristics such as rural/urban/peri-urban location, mental healthcare model, and facility level^29^.

### Patient and public involvement

We conducted formative focus group discussions and individual interviews with a variety of stakeholders (perinatal clients, non-specialist providers, specialist providers, and policymakers) to refine the IPMH intervention and select implementation strategies. A subset of 12 participants across all stakeholder groups were invited to join a participatory design group (PDG), a compensated group of policymakers, HCWs, lay providers and perinatal women who participated in a series of workshops to optimize IPMH. In addition to the PDG, a Community Advisory Board (CAB) will be convened at least twice annually. The CAB will include providers and community members who will give guidance if community issues arise and study findings will be shared with them to inform broader dissemination of results.

### Randomization

This is a cluster-randomized trial whose unit of randomization is the healthcare facility. We used restricted randomization, stratifying the 20 facilities into seven groups of 2-6 facilities, based on characteristics likely to influence outcomes of interest, including facility level, patient volume, location (rural, urban or peri-urban), and services offered. Randomization was done through an interactive event using an online visual simulation of randomization. A representative from each facility “spun” an online “wheel” to determine their order of assignment within stratified groups and a second wheel to determine final assignment within the randomization stratum^30^. Facilities were randomized 1:1 to receive IPMH intervention vs. enhanced standard of care.

### Intervention

The IPMH intervention consists of three combined evidence-based interventions: (1) mental health screening for all pregnant women who screen positive for PHQ-2 and GAD-2 with PHQ-9 and GAD-7, as recommended by mhGAP, (2) lay-delivered PM+ counseling for those with positive PHQ-9 or GAD-7 screening, and (3) telepsychiatry for women with severe symptoms, suicidality or no response to PM+ (Figure 2). This is a pragmatic trial, in which all intervention steps will be provided by facility staff, not study staff.

**Figure 2.**
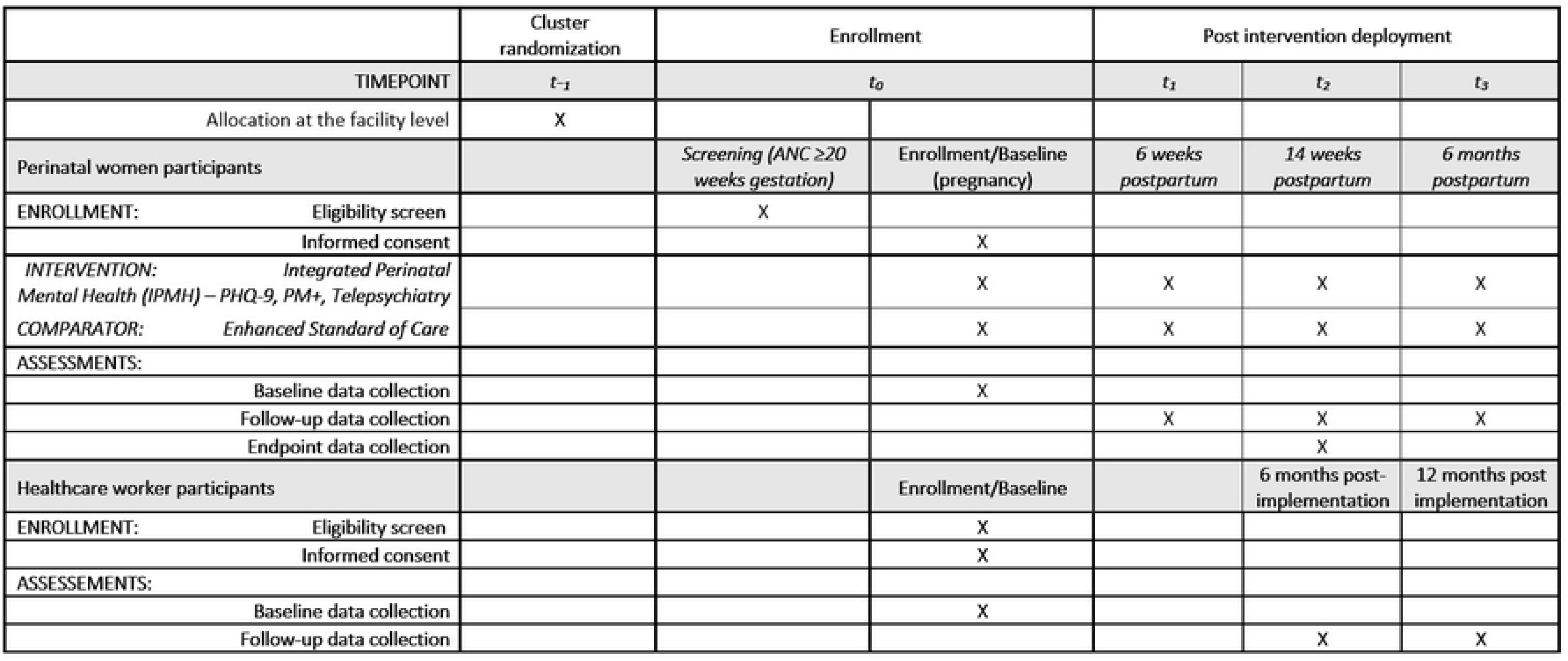
Schematic of the Integrated Perinatal Mental Health trial

#### Screening

All pregnant women attending ANC care in the intervention arm will be initially screened for PMAD symptoms by trained lay providers using the PHQ-2 and GAD-2^31,32^. MCH services in Kenya are tailored to HIV status. Women not known to be living with HIV receive HIV testing at ANC, provided by an HIV testing service (HTS) provider. Women living with HIV (WLWH) do not receive HIV testing but meet with mentor mothers (trained peers who are engaged within the healthcare system to support, educate, and empower pregnant women and new mothers living with HIV). HTS providers will administer PHQ-2 and GAD-2 screening to women living without HIV and mentor mothers to WLWH. Women with positive results (PHQ-2≥3 and/or GAD-2≥3) will be referred to the facility nurses for further assessment using PHQ-9 and GAD-7^33,34^. Those with moderate depression symptoms, defined by PHQ-9≥ 10-14 or moderate anxiety symptoms defined by GAD-7 ≥10-14, will qualify for PM+. Perinatal women with severe depression symptoms, defined as PHQ-9≥15, severe anxiety symptoms, defined as GAD-7≥15, or those who endorse suicidality (item 9 in PHQ-9) will qualify for telepsychiatry.

#### PM+

PM+ is a brief manualized transdiagnostic five-session program based on Cognitive Behavior Therapy (CBT), designed for delivery by peers or lay providers in resource constrained settings^23,24^. In this study, weekly individual PM+ sessions will be delivered in-person by trained HTS providers (for women living without HIV) and by mentor mothers (for WLWH), under the supervision of facility nurses. Perinatal women who do not respond to PM+ defined as less than 50% reduction or <5-point decrease in PHQ-9 or GAD-7 scores after completing the sessions will be offered telepsychiatry.

#### Telepsychiatry

Telepsychiatry will be provided at the study facility. Patients will connect with psychiatrists engaged by the study, through a secure Zoom platform. Intervention facilities will have a consultation room with a study-provided tablet with cellular internet to facilitate telepsychiatry sessions. The study staff will assist in setting up the telepsychiatry session. The psychiatrist will document the care plan for the patient including the diagnosis, treatment and next appointment visit if applicable in an electronic questionnaire, using REDCap^35^ and hand over to the facility nurse for next steps..

### Control treatment

Perinatal women at the control facilities will receive enhanced standard of care (eSOC) which includes initial screening for PMAD symptoms using PHQ-2 and GAD-2 by HTS providers for women living without HIV and mentor mothers for the WLWH. Perinatal women who screen positive, defined as PHQ-2 and/or GAD-2≥3, will be referred to mental health services by the nurse according to the routine practice at that facility.

### Implementation strategies

We conducted focus group discussions with women and healthcare providers, and individual interviews with policymakers to identify anticipated barriers to implementation of IPMH and potential implementation strategies to address these barriers. We then conducted a quantitative survey with perinatal clients, lay providers, non-specialist and specialist providers, to quantitatively prioritize the importance and addressability of identified barriers and implementation strategies, based on the OPTICC prioritization toolkit.^36^ Finally, we held an interactive stakeholder workshop with lay providers, non-specialist providers, psychiatrists, and policymakers where the results of the qualitative and quantitative activities were presented. During the workshop, stakeholders were invited to select and specify implementation strategies, using Proctor’s strategy specification recommendations^37^.

The following implementation strategies were identified and will be applied as outlined in the Proctor specification table^37^ (Table 1): 1) Regular audit of each facility’s performance of screening, referral, and PM+ provision, summarized and presented to facility staff to identify bottlenecks and propose actionable solutions to improve processes and outcomes; 2) Structured educational health talks for patients on perinatal mental health, provided by facility staff using prepared scripts and scenarios presented in a visual flipbook; 3) Posters displayed on walls to convey mental health information to patients; 4) Provision of essential medication, the antidepressant sertraline 50mg tablets, at all study facilities, to address medication stockouts and ensure medication availability if prescriptions are needed; 5) Financial compensation to lay providers, psychiatrists and nurses for provision of PM+, telepsychiatry and supervision sessions; 6) Structured group supervision sessions regularly offered by facility nurses to lay providers; 7) Using specifically designed paper screening forms with carbon copies to document screening results for perinatal women; 8) Quarterly interdisciplinary trainings conducted by a panel of subject matter experts, guest speakers and facility staff, each session including a didactic portion on a perinatal mental health topic, followed by a case presentation and discussion; and 9) Process mapping workflows related to each treatment step in IPMH by facility staff.

**Table 1:**
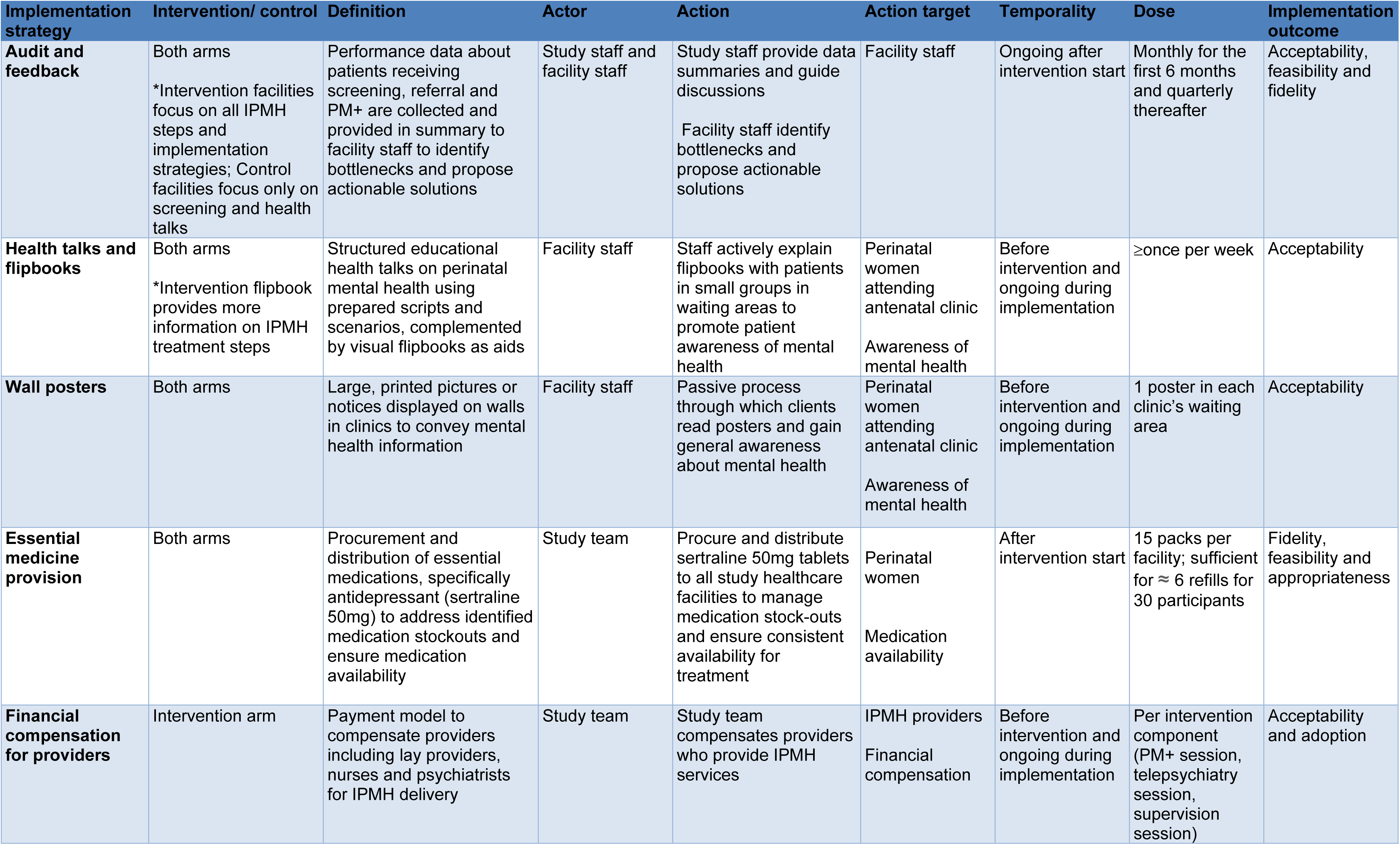
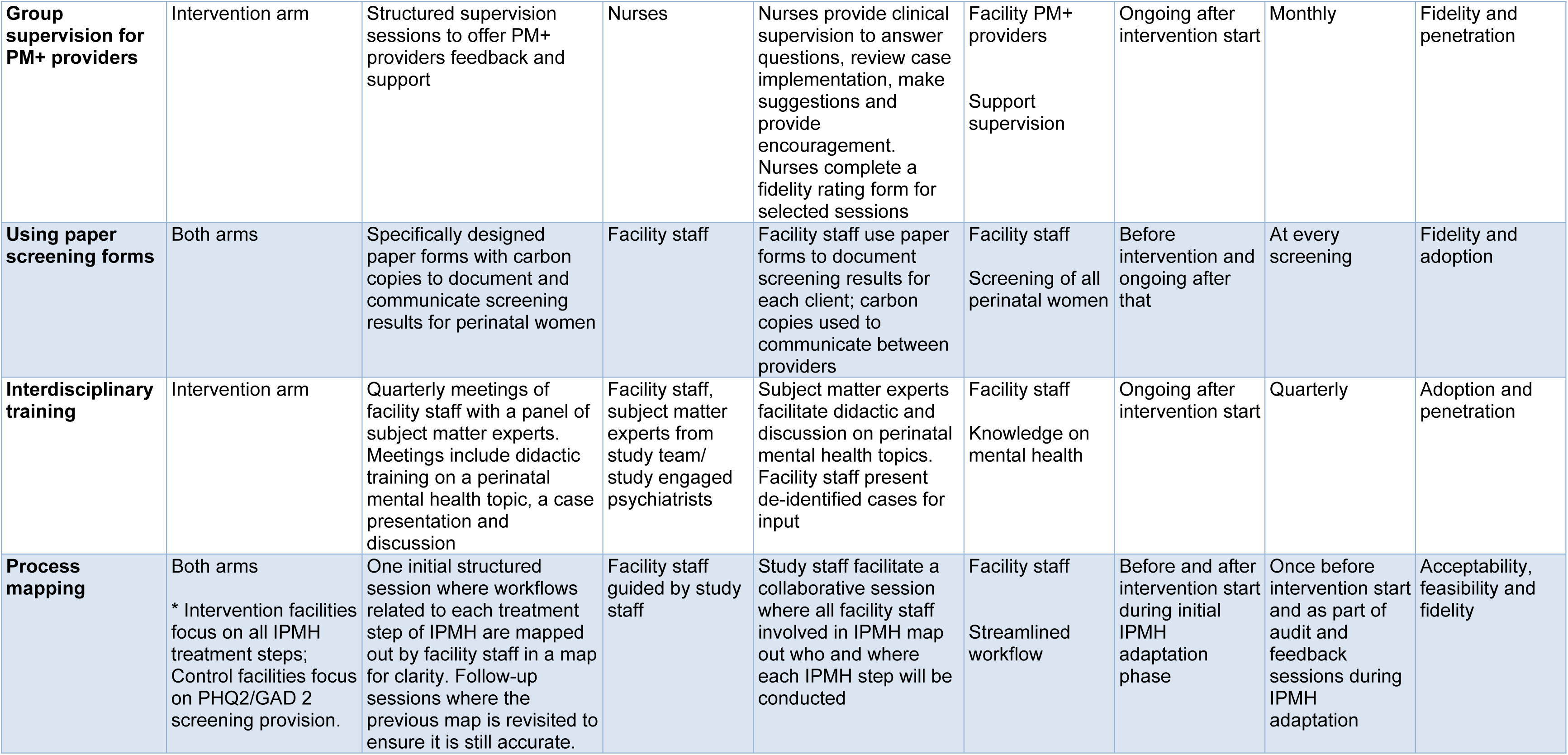
Proctor Specification Table of Implementation Strategies for Integrated Perinatal Mental Health (IPMH)

### Recruitment and screening

Study data collection will be conducted among two participant populations: perinatal clients and facility healthcare providers.

#### Perinatal clients

Study participants will be recruited from the MCH and PMTCT clinics at our study facilities. To identify potential study participants across both study arms, facility-based HTS providers and mentor mothers at all study sites will screen pregnant women attending ANC for PMAD symptoms using PHQ-2 and GAD-2. Women will be eligible for the study if they are pregnant, ≥20 weeks gestation, attending ANC at the facility, ≥14 years old and screen positive for PMAD symptoms (PHQ-2≥3 and/or GAD-2≥3). Participants on antidepressants will be eligible if they have been on stable treatment for ≥6 months. In Kenya, pregnant women below 18 years of age are considered emancipated minors and can consent to study participation independently. Women identified as high risk for self-harm based on a study self-harm assessment protocol, those with cognitive impairments, or those with psychotic symptoms will not be eligible to participate but will instead be referred for routine services at the facility including specialist care.

Pregnant women who screen positive for PMAD symptoms (PHQ-2≥3 and/or GAD-2≥3) will be referred by the facility nurse to the study staff at the conclusion of their routine facility visit for eligibility screening. Study staff will inform them about the study. If the woman is interested, study staff will administer a tablet-based screening questionnaire to assess eligibility. Eligible participants will provide written informed consent for enrolment.

#### Healthcare providers

Healthcare providers will be recruited from all cadres potentially involved in the delivery of IPMH services at all study facilities. Eligible providers include facility-in-charges, nurses, mentor mothers, and HTS providers. Study staff will collaborate with facility leadership to identify eligible providers. Following initial identification, study staff will approach providers, share information about the study, and assess interest in participation. Interested providers will undergo eligibility screening and, if eligible, will be invited to provide written informed consent for enrolment. Participation will be voluntary, and providers may withdraw at any time without consequence to their employment or clinical responsibilities.

### Enrollment and data collection

#### Perinatal clients

After obtaining informed consent, data will be collected by study staff through tablet-based questionnaires. In both intervention and control arms, enrolment data will capture sociodemographic characteristics, medical and obstetric history, and, for WLWH, details of HIV care and treatment including adherence using the Wilson 3-item self-report scale^38^. Mental health will be assessed using PHQ-9^34^ for depressive symptoms, GAD-7^33^ for anxiety symptoms, and the DSM-5 cross-cutting symptom measure to evaluate a broad range of psychiatric symptoms^39^. Intimate partner violence (IPV) will be assessed using Hurt, Insult, Threaten, Scream (HITS) scale^40^. Additional instruments will include the Reducing Tension checklist to evaluate the mechanism for PM+^41^, the Brief WHO Quality of Life (WHOQOL-BREF) to assess quality of life^42^, the Medical Outcomes Study (MOS) Social Support Survey^43^, experiences with the mental health services using acceptability (AIM), feasibility (FIM) and appropriateness (IAM) questions^44^ and mental health service utilization. At the end of the enrollment visit, participants referred to either PM+ or telepsychiatry will be scheduled for these interventions. Only the facility-administered PHQ-9 and GAD-7 will be used to determine eligibility for PM+/telepsychiatry even if study-administered tools show different treatment eligibility status.

#### Healthcare providers

Following informed consent, healthcare providers will complete a tablet-based questionnaire administered by study staff. The questionnaire will collect sociodemographic and professional background information, including provider cadre, years of experience, and current roles in IPMH service delivery. At baseline (pre-RCT launch), different implementation determinants will be assessed in intervention facilities using validated scales: Organizational Readiness for Implementing Change (ORIC)^45^, Implementation Climate Scale, Measure of Innovation-Specific Implementation Intentions (MISII)^46^, Clinician Associative Stigma Scale^47^, and measures of acceptability, feasibility and appropriateness^44^. In control facilities providers will complete the ORIC, Implementation Climate Scale, and Clinician Associative Stigma at enrollment.

### Follow-up visit

#### Perinatal clients

Follow-up study visits will be conducted by study staff at 6 weeks, 14 weeks, and 6 months postpartum in both study arms, aligned with routine postpartum and well-baby visits. At each study visit, questionnaires will be administered to assess depression symptoms using the PHQ-9^34^ and anxiety symptoms using GAD-7^33^; quality of life using WHOQOL-BREF ^42^; IPV using HITS ^40^; Reducing Tension Checklist to assess mechanism of action of PM+ ^41^; Parental Stress Scale^48^, experiences with the mental health services using acceptability (AIM), feasibility (FIM) and appropriateness (IAM) questions^44^, mental health service utilization, and adherence to antiretroviral medication for WLWH using Wilson 3-item self-report scale^38^. Pregnancy and infant outcome data will also be collected from patient facility records, the MCH booklet and from women’s self-reports. These data will include gestational age at birth, place and mode of delivery, infant sex, birth weight, infant HIV status, and whether infant HIV prophylaxis was given. Participants who miss their visits will be actively traced by phone call and/or home visit to maximize retention.

#### Healthcare providers

Serial cross-sectional data collected at 6 and 12 months after study launch at intervention facilities will include inner setting measures from the Consolidated Framework for Implementation Research (CFIR)^49,50^, Normalization Measurement Development (NoMAD)^51^ to assess sustainability, Clinician Associative Stigma Scale^47^, and measures of acceptability, feasibility and appropriateness^44^. In control facilities, providers will complete the Clinician Associative Stigma Scale at 6 and 12 months, and measures of inner setting culture and stress at 6 months.

### Clinical Outcomes

The primary outcomes are depression and anxiety symptoms, ascertained at 14 weeks postpartum by PHQ-9 and GAD-7 score change respectively. Secondary outcomes include self-reported quality of life, ascertained at 14 weeks postpartum using the WHOQOL BREF instrument; any adverse pregnancy outcome (defined as any of pregnancy loss, stillbirth, preterm birth, low birthweight, intrauterine growth restriction, neonatal hospital admission, or neonatal death at 6 weeks postpartum per participant self-report); and PM+ mechanism of action defined as score on the 10-item Reducing Tensions Checklist score at 6 weeks postpartum. Table 2 summarizes clinical outcomes.

**Table 2:**
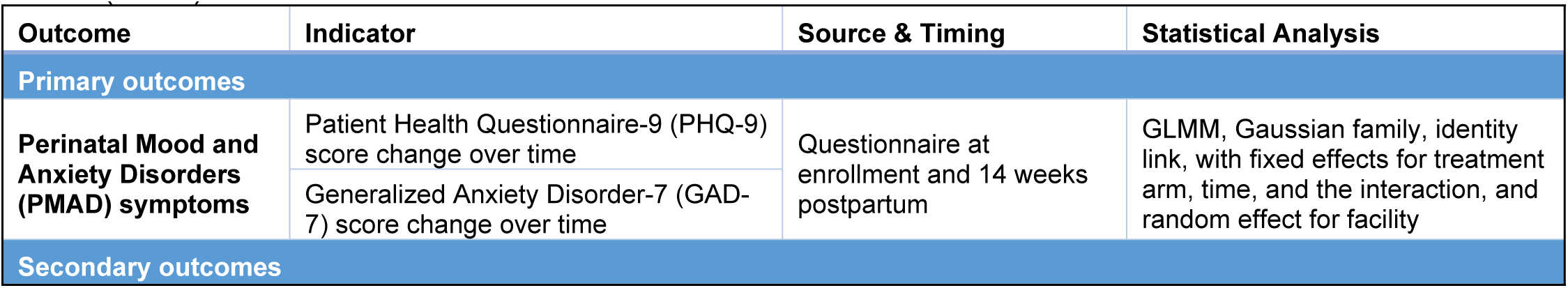
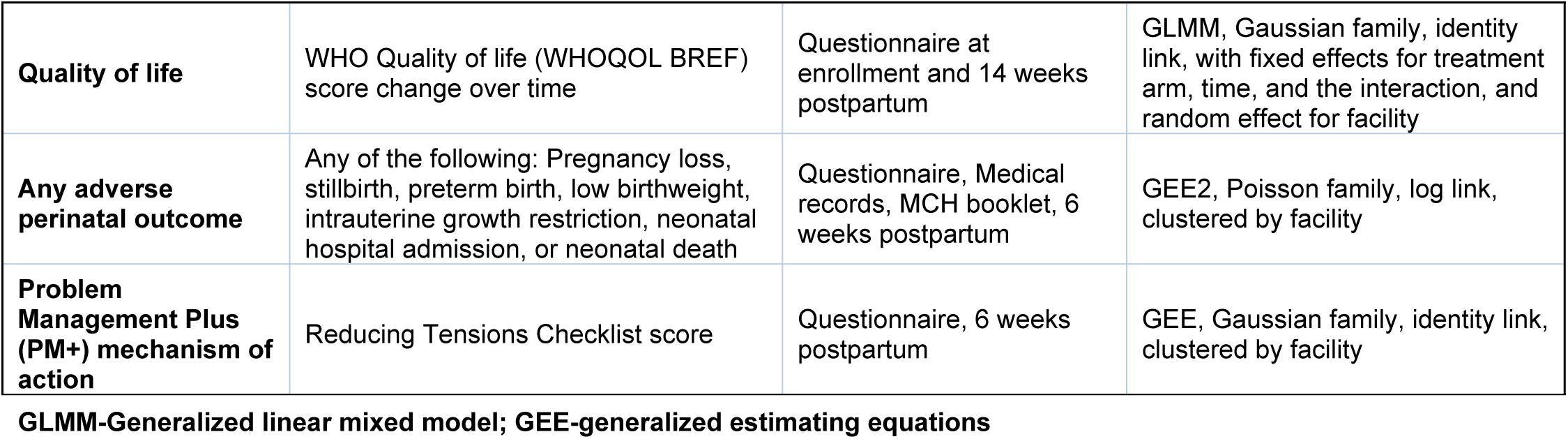
Summary of Clinical Outcomes and Analytical Approaches for Integrated Perinatal Mental Health (IPMH)

| Outcome | Indicator | Source & Timing | Statistical Analysis |
| --- | --- | --- | --- |
| Primary outcomes |  |  |  |
| Perinatal Mood and Anxiety Disorders (PMAD) symptoms | Patient Health Questionnaire-9 (PHQ-9) score change over time | Questionnaire at enrollment and 14 weeks postpartum | GLMM, Gaussian family, identity link, with fixed effects for treatment arm, time, and the interaction, and random effect for facility |
|  | Generalized Anxiety Disorder-7 (GAD-7) score change over time |  |  |
| Secondary outcomes |  |  |  |
| **Quality of life** | WHO Quality of life (WHOQOL BREF) score change over time | Questionnaire at enrollment and 14 weeks postpartum | GLMM, Gaussian family, identity link, with fixed effects for treatment arm, time, and the interaction, and random effect for facility |
| **Any adverse perinatal outcome** | Any of the following: Pregnancy loss, stillbirth, preterm birth, low birthweight, intrauterine growth restriction, neonatal hospital admission, or neonatal death | Questionnaire, Medical records, MCH booklet, 6 weeks postpartum | GEE2, Poisson family, log link, clustered by facility |
| **Problem Management Plus (PM+) mechanism of action** | Reducing Tensions Checklist score | Questionnaire, 6 weeks postpartum | GEE, Gaussian family, identity link, clustered by facility |

GLMM-Generalized linear mixed model; GEE-generalized estimating equations

### Service delivery and implementation outcomes

We will evaluate service delivery and implementation outcomes at the 20 study facilities during the implementation period. Guided by Proctor’s Implementation Outcomes Framework (IOF)^37^ and the Consolidated Framework for Implementation Research (CFIR)^49^, we will use a combination of quantitative and qualitative approaches to assess service delivery and implementation outcomes of IPMH (penetration, equity, efficiency, acceptability, fidelity and cost) and identify their multilevel drivers (Table 3).

**Table 3:** Summary of Service Delivery and Implementation Outcome Indicators and Analyses for Integrated Perinatal Mental Health (IPMH)

| Outcome | Definition | Indicator | Data source | Timepoint | Analysis |
| --- | --- | --- | --- | --- | --- |
| **Compare control vs intervention arms** |  |  |  |  |  |
| **Penetration** | Extent to which MCH and PMTCT clients receive screening & treatment for PMAD | - Proportion of clients receiving recommended screening (PHQ-9 and GAD-7)<br>- Proportion of positive screeners linking to any mental healthcare | - Perinatal women self-report questionnaires<br>- Perinatal women self-report questionnaires | Cross-sectionally per client throughout study | ME binomial regression (RE by facility) |
| **Equity** | Absence of avoidable differences in care delivery between groups of people | - Disparities in accessing: screening<br>- any mental healthcare between quintiles of demographic characteristics (e.g., age, income, distance to facility) | - Perinatal women self-report questionnaires | Cross-sectionally per client throughout study | ME linear or binomial regression (RE by facility); intervention arm as interaction term |
| **Efficiency** | Minimize intervention and implementation strategy inputs for number of perinatal women receiving care. | - Average number of provider hours per month per perinatal woman<br>- Average number of hours perinatal women wait time per month<br>- Average incremental cost per perinatal women screened & treated (see cost) | - Time & motion study; direct observation of patient flow and provider time<br>- Routine program data abstraction | - RCT launch<br>- 6 months post-launch<br>- 12 months post-launch | ME linear regression (RE by facility) |
| **Cost** | Incremental costs to health system & client from IPMH implementation | - Average incremental total facility cost by arm<br>- Average incremental unit cost per facility by arm, where unit costs are defined as incremental cost per client screened or per client screened and treated | - Activity-based costing<br>- Time and motion of provider time and non-personnel resource use<br>- HCW & perinatal women out of pocket questionnaire, IDI or FGD<br>- Project and MOH facility expense reports | - RCT launch<br>- 12 months post-launch |  |
| **Evaluate within intervention arm** |  |  |  |  |  |
| **Acceptability** | Perception that IPMH is agreeable or satisfactory | - Acceptability of Intervention Measure (AIM) score (Perinatal & HCWs)<sup>52</sup><br>- Qualitative assessment | - HCW & perinatal women questionnaires & IDIs | - RCT launch<br>- 6 months post-launch<br>- 12 months post-launch | - Descriptive quantitative & qualitative<br>- Quantitative determinants by ME linear regression (RE by facility, RE by perinatal women/HCW respondent for acceptability & fidelity)<br>- Qualitative determinants by thematic analysis |
| **Fidelity** | The extent to which IPMH is implemented as designed | - Proportion of positive screeners receiving 5 sessions of PM+<br>- Proportion of positive screeners receiving all PM+ manual components<br>- Proportion of PM+ non-responders receiving tele-linkage<br>- Lay provider competence in PM+ following initial training | - Perinatal women report of PM+ services delivered and provider logs<br>- Self-administered HCW competence checklist based on content of training | - 6 months post-launch<br>- 12 months post-launch |  |

ME- Marginal Effects

Penetration will be defined as the proportion of clients receiving the recommended screening and treatment, based on provider logs and self-reported data from perinatal women. Equity will be defined as the absence of differences in screening and treatment rates among groups with different demographic characteristics. We will compare rates of screening and any mental healthcare services provided between quintiles of demographic characteristics (e.g., age, income, distance to facility). Efficiency will be defined as time and cost spent on regular clinical care (perinatal visit completion, overall HCW workload). We will collect time-and-motion data on patient flow to estimate client wait times, provider workload, and resource use. Fidelity will be assessed by measuring adherence to the PM+ protocol, including the number of sessions received, delivery of all core components, and referral of non-responders for telepsychiatry, based on provider activity logs. Acceptability will be measured using the Acceptability of Intervention Measure (AIM) completed by both perinatal women and healthcare workers (HCWs). In addition, we will conduct a qualitative evaluation, as described below, using in-depth interviews to further explore satisfaction, perceived value, and fidelity.

#### Recruitment and data collection for qualitative evaluation of IPMH acceptability and fidelity

We will use purposive sampling to recruit perinatal women who receive PM+ and/or telepsychiatry and HCWs involved in IPMH delivery (HTS providers, lay HCWs, facility nurses, and psychiatrists) to participate in in-depth interviews (IDIs) at 6 month and 12 month post-trial launch. Eligible perinatal women will be ≥18 years and sampling will seek variation across key implementation scenarios: completion of PM+, delays or restarts, non-response to PM+ with escalation to telepsychiatry and direct referral to telepsychiatry. Participants will be identified via clinic registers and staff referral, approached in person or by phone, and invited to opt in. We will conduct10-15 IDIs with perinatal women and 15-20 IDIs with HCWs, stratified by cadre, until thematic saturation is reached (operationalized as no new codes/ themes across two consecutive interviews within a stakeholder group). Trained qualitative researchers not involved in clinical care will conduct interviews in private rooms at facilities or neutral venues. IDIs will explore participant’s overall experiences, acceptability, and fidelity of IPMH intervention as well as contextual factors that may influence its delivery and uptake. Sessions will last ∼45-60 minutes and follow semi- structured guides aligned with CFIR using contextually adapted questions.

With written informed consent, interviews will be audio-recorded, and conducted in English, Kiswahili or Dholuo. Recordings will be transcribed verbatim and, if needed, translated into English. A team of at least two analysts will develop a codebook using a combination of deductive and inductive methods. Deductive codes will be drawn from the CFIR pre-developed codebook. Qualitative and quantitative data will be combined in a convergent mixed methods approach for triangulation.

#### Costing

We will conduct a prospective micro-costing study using expenditures and activity-based costing to estimate the economic costs of the intervention and its implementation from a societal perspective. We will obtain expenditure data directly from project records or facility financial records to ensure that we capture all labor (personnel time, volunteer labor) and non-labor costs, such as commodities, capital goods (e.g., vehicles, computers, cell phones, other equipment, overhead), costs associated with IPMH development and delivery.

We will conduct time and motion studies to estimate both health system and patient resource use. For resource use not captured in project expense reports, we will conduct interviews with IPMH providers, perinatal women, and administrators on the resource use for each intervention component and implementation strategy, capturing key information on quantities and prices each activity, as possible. All goods and services will be valued regardless of if they were paid for by the project or donated. For perinatal women, we will estimate the value of their time traveling to and from facilities and receiving care services, as well as costs incurred for childcare and lost wages. To estimate total incremental economic and unit costs, we will estimate the cost of IPMH plus implementation strategies compared to enhanced standard of care in control facilities, along with the number of participants receiving care in each arm. We will conduct costing activities in a subset of 5-10 facilities purposively selected to represent heterogeneity in drivers of cost (e.g., patient volume, staffing, location, prevalence of PMAD symptoms). We will estimate start-up and recurrent costs for all intervention and relevant implementation strategies following best practices^53^.

### Sample size

#### Perinatal women

Our study is powered for the primary outcomes of depression and anxiety symptom scores. We will enroll 2,970 participants across our 20 study facilities. The number recruited at each facility will be allowed to vary between 130 and 170. With 2,970 perinatal women, assuming α=0.05, 2-sided testing, 10 clusters per arm, a coefficient of variation of 0.25 between clusters, and unequal cluster sizes, we will have ≥80% power to detect an absolute difference of 1.66 in continuous PHQ-9 and GAD-7 score (9.50 vs. 7.84 mean PHQ-9 score and 9.00 vs. 7.34 GAD-7 score). We will also have ≥80% power to detect a difference of 0.17 in continuous WHOQOL BREF score (3.40 vs. 3.57), 11.5% in any adverse perinatal outcome among all women (35.0% vs. 23.5%), and 2.2 in continuous reducing tensions checklist score (27.0 vs. 29.2).

#### Healthcare workers

For the cross-sectional surveys conducted at baseline, 6-month post-implementation, and 12-month post-implementation, we will enroll approximately 200 healthcare providers across the 20 study facilities. The number of providers recruited at each facility will vary between 8 and 12, depending on staffing levels and roles in IPMH delivery. While the sample size will not provide sufficient power to detect statistically significant changes in implementation determinants or outcomes, it will allow us to descriptively assess patterns and trends over time.

### Statistical analysis

The distribution of baseline characteristics both pooled and by study arm will be described using summary statistics appropriate for the measurement scale. To determine if randomization is balanced, we will compare baseline characteristics between study arms using Chi-squared tests for categorical variables and Kruskal-Wallis tests for continuous variables. Any variables found to vary significantly at baseline will be included as adjustments in the primary analysis. Using an intent-to-treat analysis, change in PHQ-9, GAD-7 and WHOQOL BREF scores from enrollment to 14 weeks postpartum will be compared between arms using generalized linear mixed models (GLMM) with a Gaussian family. We will include fixed effects for treatment arm, time since enrollment, and the interaction between treatment and time, and random effect for facility (Table 2). The proportion of women with any adverse pregnancy outcome will be compared between arms using GEE with a Poisson family, exchangeable correlation structure and robust errors, clustered by facility. Reducing Tensions Checklist score at 6 weeks postpartum will be compare between arms using GEE with a Gaussian family and exchangeable correlation structure, clustered by facility.

We will repeat analyses for PMAD symptoms and Quality of Life at 6 weeks postpartum and 6 months postpartum as pre-specified exploratory analyses. The 6-week postpartum analysis will be conducted to evaluate whether a more proximal time point to the conclusion of PM+ engagement detects a larger effect. The 6-month postpartum analysis will be conducted to evaluate whether an effect detected at 14 weeks postpartum is sustained into later postpartum. Additionally, to evaluate differences in trajectory of symptoms between study arms, we will conduct exploratory analyses that include symptom scores from all follow-up visits (enrollment, 6 weeks, 14 weeks, 6 months postpartum). We will fit GLMM with Gaussian family, with fixed effects for treatment arm, time, and an interaction term for arm and time, and a random effect for facility. We will perform 3 separate models for the relevant outcomes (PHQ-9 score, GAD-7 score, WHOQOL BREF score).

We will conduct subgroup analyses of primary and secondary outcomes by HIV status. We anticipate 10-15% of our population will be living with HIV. Assuming alpha=0.05, 2-sided test, 10 clusters per arm, N=405, unequal cluster sizes, 10% attrition, and a conservative coefficient of variation between clusters=0.25, we will have ≥80% power to detect a difference in continuous PHQ-9 and GAD-7 score among WLWH of 2.15: 9.5 vs. 7.35 mean PHQ-9 score and 9 vs. 6.85 GAD-7 score. We will have ≥80% power to detect a difference in continuous WHOQOL BREF score of 0.22 among WLWH: 3.4 vs. 3.62 mean score.

To evaluate implementation outcomes, we will conduct both between-arm and within-arm comparisons of implementation outcomes using appropriate GLMM models to account for clustering at the facility level. Binary outcomes, such as receipt of mental health screening and linkage to care, will be analyzed using GLMM with binomial family, while continuous measures, such as wait times, will be assessed using GLMM with Gaussian family. Facility will be specified as a random effect in all models, and where outcomes are measured at the individual level (e.g., acceptability), additional random effects will be included for respondent (perinatal woman or healthcare worker). Equity will be analyzed and compared between and within arms by testing for differences in care delivery across demographic subgroups by binomial regression. The effect of intervention arm on associations with demographic characteristics will be evaluated by testing for effect modification by incorporating an interaction term in the GLMM. Efficiency outcomes, including client wait time and provider time allocation, will be assessed using data from time-and-motion studies. Acceptability and fidelity will be described quantitatively and qualitatively, with scores modeled as a function of facility and individual-level determinants. Thematic analysis of interview data using CFIR will further explore perceptions of implementation success and contextual influences. Cost analyses will apply activity-based costing (Table 3).

## Discussion

This study will be among the first trials to evaluate a stepped care approach to integrate perinatal mental health services into routine public MCH and PMTCT clinics in a low resource setting, incorporating telepsychiatry as part of the care model. This multi-component approach combining screening, psychotherapy delivered by lay providers and in-facility tele-psychiatry is novel and aligns with the WHO recommendation and Kenya’s national mental health strategy. The intervention seeks to address critical barriers to delivery of PMAD services to perinatal women who need it the most without straining the overburdened mental health care delivery system. Optimization of the intervention together with incorporation of implementation strategies derived by engagement of multiple stakeholders enhances feasibility, acceptability and the likelihood of widespread adoption and scalability of the approach. By including both clinical effectiveness and implementation outcomes, the findings from this trial have strong potential to inform policy decisions and clinical guidelines for the care of perinatal women.

### Current study status

Participant enrolment began on 17^th^ February 2025. We anticipate that recruitment will be complete on 28^th^ February 2027 and data collection will be completed 31^st^ March 2028. Results are expected in December 2028. Recruitment, intervention delivery and data collection are currently ongoing.

### Ethics and dissemination

The study has received ethical approval at the Kenyatta National Hospital/ University of Nairobi (P425/04/2023) and the University of Washington (STUDY00017933) institutional review boards. All participants will provide written informed consent prior to enrollment. Upon enrollment, each participant is assigned a unique study identification number, which is used in all study records to maintain confidentiality. Personal, identifying information is stored in a secure, password-protected, encrypted REDCap database, separate from the main study datasets. All study personnel are trained in human subjects, research ethics and study-specific standard operating procedures. Study findings will be disseminated to key stakeholders, including policymakers at county and national levels in Kenya, as well as the research community, through peer-reviewed publications and presentations at scientific conferences. Cleaned, de-identified datasets will be made available within 12 months of publication of the primary study findings.

## Data Availability

No datasets were generated or analysed during the current study. All relevant data from this study will be made available upon study completion.

## Authors’ contributions

JK, KR, and AB conceived the study and designed the protocol, and provided oversight and guidance on study design and methodology. AKK and NG drafted the first version of the manuscript. ADW, AL, KR, AB, LO, and AR developed study and training materials. BAR and KR developed the study analysis plan. DO, AL, YW, and LP contributed to database development. SM provided guidance on costing. AKK, NG, AN, AK, CM, and CN contributed to implementation planning. All authors reviewed, revised, and approved the final manuscript.

